# A randomized double-blind placebo-controlled trial of Guanfacine Extended Release for aggression and self-injurious behavior associated with Prader-Willi Syndrome

**DOI:** 10.1101/2024.08.22.24312419

**Authors:** Deepan Singh, Michael Silver, Theresa Jacob

**Affiliations:** Department of Psychiatry, Maimonides Medical Center, Brooklyn, New York; Research Administration, Maimonides Medical Center, Brooklyn, New York; Clinical and Translational Research Labs, Maimonides Medical Center, Brooklyn, New York

## Abstract

**Introduction:** Prader-Willi Syndrome (PWS), a rare genetic disorder, affects development and behavior, frequently resulting in self-injury, aggression, hyperphagia, oppositional behavior, impulsivity and over-activity causing significant morbidity. Currently, limited therapeutic options are available to manage these neuropsychiatric manifestations. The aim of this clinical trial was to assess the efficacy of guanfacine-extended release (GXR) in reducing aggression and self-injury in individuals with PWS.

**Trial Design:** Randomized, double-blind, placebo-controlled trial conducted under IRB approval.

**Methods:** Subjects with a diagnosis of PWS, 6-35 years of age, with moderate to severe aggressive and/or self-injurious behavior as determined by the Clinical Global Impression (CGI)-Severity scale, were included in an 8-week double-blind, placebo-controlled, fixed-flexible dose clinical trial of GXR, that was followed by an 8-week open-label extension phase. Validated behavioral instruments and physician assessments measured the efficacy of GXR treatment, its safety and tolerability.

**Results:** GXR was effective in reducing aggression/agitation and hyperactivity/noncompliance as measured by the Aberrant Behavior Checklist (ABC) scales (p=0.03). Overall aberrant behavior scores significantly reduced in the GXR arm. Aggression as measured by the Modified Overt Aggression Scale (MOAS) also showed a significant reduction. Skin-picking lesions as measured by the Self Injury Trauma (SIT) scale decreased in response to GXR. No serious adverse events were experienced by any of the study participants. Fatigue /sedation was the only adverse event significantly associated with GXR. The GXR group demonstrated significant overall clinical improvement as measured by the CGI-Improvement (CGI-I) scale. (p<0.01).

**Conclusion:** Findings of this pragmatic trial strongly support the use of GXR for treatment of aggression, skin picking, and hyperactivity in children, adolescents, and adults with PWS.

**Trial Registration:** ClinicalTrials.gov Identifier: NCT05657860

## Introduction

Prader-Willi syndrome (PWS) is a rare, complex multisystem genetic disorder, which includes hypothalamic dysfunction, cognitive and behavioral problems, increased anxiety, and compulsive behaviors. ^1, 2^ It is the most common genetic cause of morbid obesity in children. ^2^ While hyperphagia is seen as the *sine qua non* of PWS, other behavioral symptoms such as aggression, oppositional behavior, and temper tantrums, are common and cause significant distress to patients and caregivers. ^3, 4, 5^ PWS also has a high prevalence of self-injurious behavior such as skin-picking, as well as repetitive motor activity, impulsivity, and hyperactivity.^6, 7, 8, 9^ Despite the high prevalence of behavioral disturbances in PWS, evidence-based pharmacological treatment options for them remain limited. ^10^ In fact, at the time of this writing, there are no published results from any randomized controlled clinical trials evaluating the efficacy of any agent against the common behavioral disturbances seen in PWS except for hyperphagia. This is particularly concerning given that close to half of all children between ages 11-17 and over 65% of all adults with PWS are on at least one prescribed psychotropic medication with most of them requiring more than one medication.^11^

Currently, serotonin reuptake inhibitors (SRIs) and atypical antipsychotics are the most frequently utilized medications to manage behavioral problems in PWS. ^11^ Both these classes of medications are associated with the potential for developing or worsening metabolic syndrome. ^12, 13, 14, 15^ Due to the high risk of obesity in PWS, it is important to avoid weight gain-inducing agents as much as possible. Furthermore, there have been reports of the development of psychotic symptoms such as auditory/visual hallucinations and paranoid delusions with the use of SRIs and serotonin-agonists, making serotonergic agents potentially dangerous in this population. ^16, 17^ Impulsivity and hyperactivity are well-known presenting signs of attention-deficit/hyperactivity disorder (ADHD), and as many as 25% of the patients with PWS exhibit these symptoms.^7^ Stimulants for the management of impulsivity and hyperactivity may sometimes worsen mood, and irritability, and chronic use of stimulants may lead to an increase in skin-picking behavior. ^18^ Although N-acetyl cysteine and oxytocin have been suggested for the management of skin-picking behaviors, safe and effective modalities remain sparse for the management of behavioral symptoms in PWS. ^19, 20, 21, 1^

Guanfacine extended-release (GXR) is a postsynaptic α2-adrenoceptor agonist approved by the Food and Drug Administration (FDA) for the management of ADHD. ^22, 23^ Guanfacine is proposed to work by moderating the activity of the left dorsolateral prefrontal cortex associated with the emotional biasing of response execution, thus reducing impulsivity. ^24^ Medications that operate as α2-adrenoceptor agonists such as guanfacine are beneficial in treating disruptive and aggressive behaviors. ^25^ In addition, GXR has been demonstrated to be safe and effective for reducing hyperactivity, impulsivity, and distractibility in patients with autism. ^26, 27^ Most importantly, GXR has had no reports of significant weight gain, worsening hyperphagia, or other metabolic side effects. ^28, 29^ A retrospective cohort study has demonstrated that GXR led to improvement in symptoms of skin picking, aggression/agitation, and ADHD in patients with PWS.^30^ Given the significant morbidity caused by these behavioral problems in this already vulnerable population, novel evidence-based approaches are warranted for the management of aggression, impulsivity, and self-injury in PWS. ^31^ It was postulated that GXR is a safer treatment option for self-injurious and aggressive behaviors in PWS and we wanted to test this with a pragmatic approach.

The objective of this randomized, double-blind, placebo-controlled trial was to determine the efficacy of guanfacine extended release (GXR) in reducing aggression and self-injury in individuals with PWS, and to assess its safety and tolerability.

## Methods

### Study Design

This pragmatic two-phase 16-week study was designed to test the hypothesis that GXR is effective in reducing aggression and self-injury in individuals with PWS with moderate to severe aggressive and/or self-injurious behavior. In the first phase, the randomized controlled trial part, eligible subjects were randomly assigned in a 1:1 ratio, without stratification, to GXR or placebo for 8 weeks. This was the double-blind, placebo-controlled, fixed-flexible dose phase of the clinical trial. Immediately following the blinded randomized trial period, an 8-week open-label extension was pursued to further define the efficacy and tolerability of GXR, as well as to establish its safety with a specific focus on metabolic profile.

The study was performed in accordance with all current applicable regulations, the International Conference on Harmonization of Good Clinical Practice, the principles of the Declaration of Helsinki and local ethical and legal requirements. It was approved by the Institutional Review Board (# 2020-11-03-MMC) and registered at ClinicalTrials.gov (identifier: NCT05657860). It was conducted in a mental health clinic setting of a large, urban, community-based teaching hospital in a major metropolis of the United States. The study recruitment period began March 1, 2021 and ended October 31, 2023. Study participants received complete information regarding the protocol during the consent process before enrollment. Written, IRB-approved informed consent was obtained from each participant’s parent or legal guardian, and assent was obtained from each participant, as applicable.

### Inclusion/Exclusion Criteria

To be eligible for inclusion, subjects had to be between 6 and 35 years of age with a diagnosis of PWS confirmed by genetic testing. In addition, a rating of moderate or above on the Clinical Global Impression - Severity scale (CGI-S) was required for inclusion. Females of childbearing potential had to have a negative urine pregnancy test at screening and baseline and to comply with any protocol contraceptive requirements. In addition, participants and their parent/legal guardian had to be willing, able, and likely to fully comply with the study procedures and restrictions defined in the protocol. Exclusionary criteria included a positive pregnancy test, swallowing difficulty, or the presence of clinically significant bradycardia or hypotension. Those currently on GXR, actively enrolled in other studies, or with a lactose intolerance or allergy were also excluded. Subjects receiving antipsychotic medications due to a documented history of psychosis or bipolar disorder were allowed to continue taking the medication without dosage modification. The pragmatic approach of this trial also allowed growth hormone, thyroid hormone replacement treatment, and non-psychiatric medicines, to continue albeit without dosage modifications. Similarly, N-Acetyl Cysteine (NAC) and anticonvulsant medications if prescribed for seizures were allowed, with specific instructions to refrain from making any dosage changes during the trial period.

### Study Drug Administration

The starting dose for all subjects was 1 mg per day. Subjects weighing less than 25 kg remained on the 1 mg dose until day 14. If the medication was well tolerated, the dose was raised to 2 mg until day 28 and increased to 3mg for the remaining 4 weeks in the trial. Subjects weighing 25 kg or more were eligible for an increase to 2 mg at day 7, 3 mg at day 14, and 4 mg at day 21 or day 28. The dose schedule was not fixed; the treating clinician could delay a planned increase or lower the dose to manage adverse effects. Medication was dispensed at the baseline visit and at the unblinding visit for the subsequent dosing period. GXR was administered as 1 mg capsules; placebo group received matching placebo capsules.

### Randomization and Blinding

Subjects were randomly assigned to either the placebo or experimental (GXR) group in a 1:1 ratio via a computer-generated randomization scheme with randomly permuted blocks of varying sizes, done by Pharmacy. As the first phase of this study was double-blinded, the study pharmacist was the only team member aware of patient treatment assignments. Randomization occurred at baseline (visit 1). Unblinding was at week 8, and subjects continued treatment for 8 more weeks (open-label phase).

### Assessments

Baseline and follow-up assessments included the Aberrant Behavior Checklist, (ABC), Self-Injury Trauma (SIT) scale, physician-administered Clinical Global Impression – Severity (CGI-S; only at baseline), and CGI-Improvement (CGI-I) scales, Hyperphagia Questionnaire for use in PWS clinical trials (HQ-CT), Modified Overt Aggression Scale (MOAS), and the Epworth Sleepiness Scale (ESS) at biweekly study visits. The ABC includes an overall score and sub-scores to monitor irritability, lethargy, inappropriate speech, hyperactivity and stereotypies.

In addition, laboratory tests, medical and psychiatric histories, physical examination, electrocardiography (ECG), and vital signs evaluation were conducted at study visits. Daily monitoring of vitals and problematic behavior was performed by caregivers.

### Safety Monitoring

Blood pressure and pulse rate were measured at each visit. Laboratory tests were conducted at baseline and were repeated at week 8 and week 16. An ECG was obtained at baseline and at week 8 to assess the impact of GXR on cardiac conduction. Adverse events were systematically reviewed and documented. Side effects were measured using open-ended clinician inquiry with questions about drowsiness, fatigue, decreased appetite, emotional/tearful, dry mouth, irritability, anxiety, headache, increased energy, mid-sleep wakening, stomachache, constipation, increased repetitive behavior, aggression, depressed mood, cough/congestion, self-injury, nausea, trouble falling asleep, dizziness, silly, weakness, diarrhea, vomiting, increased appetite, excessive talking, blurred vision, skin rash/eczema, nightmares, enuresis, motor tics, skin-picking, and pyrexia.

Subjects who were randomly assigned to placebo group and did not show a positive response by 8 weeks, were offered open-label treatment with GXR for 8 weeks. All subjects in this continuation phase were seen every 2 weeks for monitoring. Those unblinded and started on GXR were monitored weekly for the first 4 weeks of the continuation phase.

### Data Analysis

Data were analyzed using an intent-to-treat approach. Patient baseline data were summarized via descriptive statistics such that continuous variables were summarized by means (SD) or medians (IQR) and categorical data by counts (%). Baseline factors (CGI-S, age, gender) as well as all study observations were presented within each group. In the analysis, distributions were compared between treatment groups at baseline via the Wilcoxon rank sum test. Finally, we created a repeated measures linear regression model for each outcome scale, using Generalized Estimating Equations (GEE) to account for within-subject variability, with the predictor of treatment, to assess the impact of treatment across the entire timeline of the study. An unstructured correlation matrix was assumed. All models were then plotted in a Forest Plot for ease of comparison. Two-sided p-values < 0.05 were considered statistically significant. All analyses were conducted using SAS/STAT version 9.4 (SAS Institute Inc., Cary N.C. USA).

## Results

### Demographics

Over the study period, 65 patients with a clinical diagnosis of PWS were screened. Of these, 14 underwent randomization. Participant disposition in the study is shown in the CONSORT flow diagram (Fig 1). The study population included 11 males and 3 females, aged between 6 to 35 years, carrying genetically confirmed diagnoses of PWS. Of these subjects, 78.5% were Caucasian, and 7% were Asian. Table 1 reviews the demographics of the participants. Subtype distribution showed that 29% of the subjects carried paternal deletion and 43% had maternal uniparental disomy. Further, the submitted test reports did not distinguish subtype in 28% of the participants.

**Fig 1.**
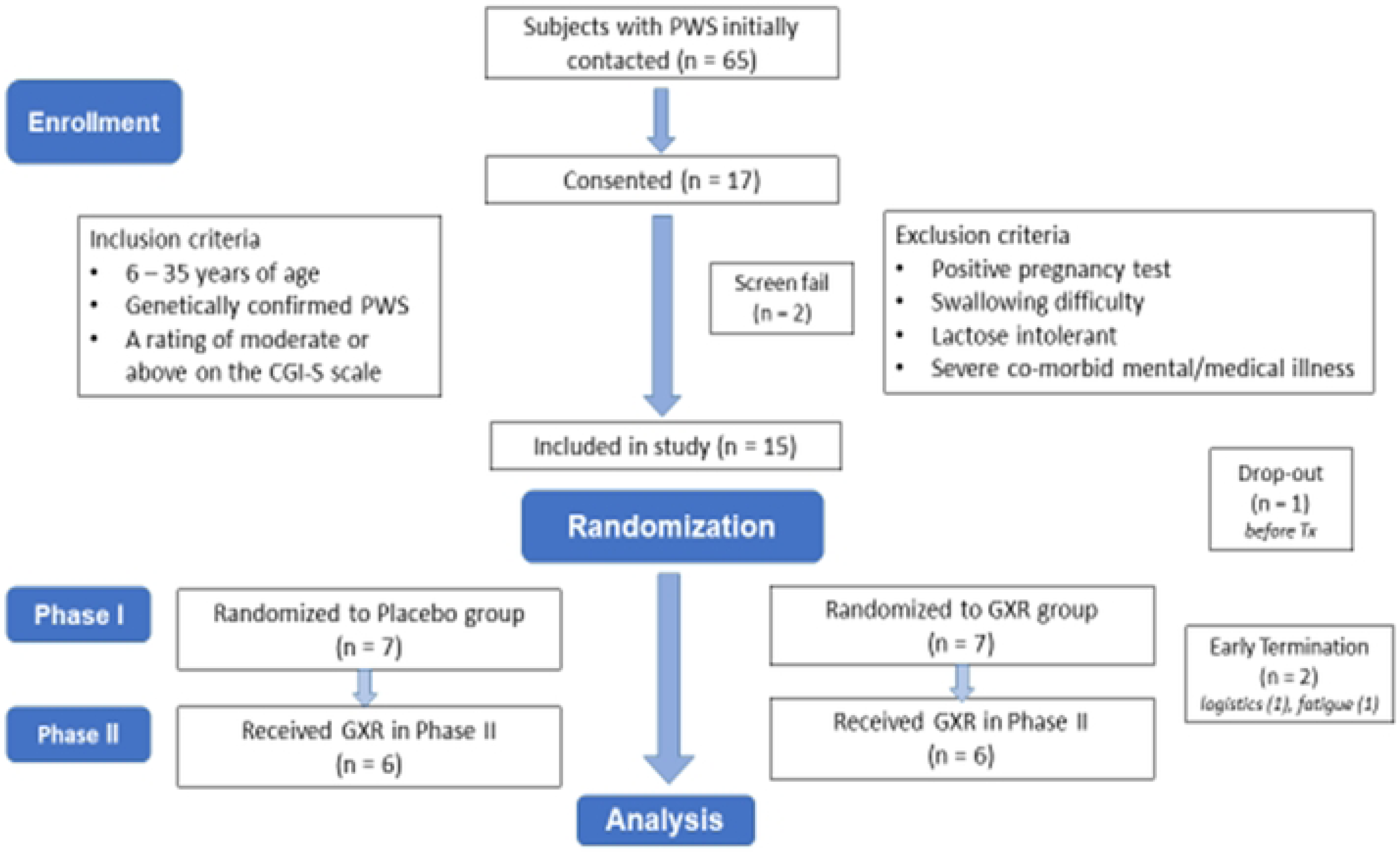
CONSORT flow diagram. Participant disposition in double-blind, randomized placebo-controlled trial for GXR (guanfacine-extended release)

**Table 1.** Baseline Demographics and Clinical Characteristics of Study Participants.

|  |  | Placebo<br>(n=7) | Guanfacine XR<br>(n=7) | P-value |
| --- | --- | --- | --- | --- |
| **Age (years)** |  | 8 (8-16) | 10 (7 - 10) | 0.383 |
| **Sex** | *Female* | 1 (14.3%) | 2 (28.6%) | 1.000 |
|  | *Male* | 6 (85.7%) | 5 (71.4%) |  |
| **Ethnicity** | *Hispanic or Latino* | 1 (14.3%) | 2 (28.6%) | 1.000 |
|  | *Not Hispanic or Latino* | 6 (85.7%) | 5 (71.4%) |  |
| **Race** | *Missing* | 1 (14.3%) | 0 (0%) | 1.000 |
|  | *Asian* | 0 (0%) | 1 (14.3%) |  |
|  | *Middle Eastern* | 0 (0%) | 1 (14.3%) |  |
|  | *White* | 6 (85.7%) | 5 (71.4%) |  |
| **Blood Pressure (mmHg)** | *Systolic* | 123 (103 - 126) | 103 (101 - 121) | 0.383 |
|  | *Diastolic* | 77 (68 - 81) | 72 (60 - 74) | 0.535 |
| **Heart Rate (bpm)** |  | 82 (63 - 101) | 87 (74 - 101) | 0.5 |
| **ECG (QTcF)** |  | 397 (384 - 413) | 398 (391 - 413) | 0.805 |
| **Height (cm)** |  | 147.32 (132.5 - 152) | 136 (124.8 - 152.4) | 0.456 |
| **Weight (kg)** |  | 65.8 (33.2 - 77.6) | 48.2 (29.8 - 66.8) | 0.318 |
| **BMI (Body Mass Index)** |  | 28.5 (18.7 - 35.2) | 23.6 (18.1 - 30.7) | 0.456 |

All numeric variables were summarized with median and IQR and compared across groups with a *Wilcoxon Rank Sum Test*. All categorical variables were summarized with frequency and percentage and compared across groups with a *Fisher Exact Test*.

### Patient Baseline Characteristics

As seen in Table 2, there were no differences among the groups at baseline for any measures, including severity of illness as measured by the CGI-S.

**Table 2.** Subject Behavioral Measure Scores at Index Visit.

| Outcome Measures | Placebo<br>(n=7) | GXR<br>(n=7) | P-value |
| --- | --- | --- | --- |
| ABC | 58 (45 - 67) | 48 (24 - 70) | 0.620 |
| ABC - Subscale I, *Irritability* | 21 (15 - 23) | 15 (6 - 26) | 0.710 |
| ABC - Subscale II, *Social Withdrawal* | 16 (5 - 20) | 11 (2 - 24) | 0.456 |
| ABC - Subscale III, *Stereotypic Behavior* | 2 (0 - 6) | 0 (0 - 2) | 0.383 |
| ABC - Subscale IV, *Hyperactivity/Noncompliance* | 16 (12 - 24) | 9 (6 - 22) | 0.209 |
| ABC - Subscale V, *Inappropriate Speech* | 5 (4 - 8) | 7 (4 - 9) | 0.710 |
| CGI-S | 6 (5 - 6) | 5 (5 - 6) | 0.535 |
| ESS | 10 (6 - 13) | 7 (1 - 8) | 0.383 |
| HQ-CT | 14 (8 - 16) | 14 (6 - 25) | 0.805 |
| MOAS (total weighted score) | 11 (11 - 17) | 13 (9 - 17) | 1.000 |
| MOAS - *Verbal Aggression Subscale* | 2 (1 - 2) | 2 (2 - 2) | 0.805 |
| MOAS - *Aggression Against Property Subscale* | 2 (2 - 4) | 2 (0 - 4) | 0.620 |
| MOAS - *Autoaggression Subscale* | 3 (3 - 3) | 3 (3 - 6) | 0.259 |
| MOAS - *Physical Aggression Subscale* | 4 (3 - 8) | 8 (0 - 8) | 0.902 |
| SIT - *Number Index Score* | 1 (1 - 2) | 2 (1 - 2) | 0.620 |
| SIT - *Severity Index Score* | 2 (1 - 5) | 2 (1 - 3) | 1.000 |

**ABC**, Aberrant Behavior Checklist; **CGI-S**, Clinical Global Impression – Severity scale; **ESS**, Epworth Sleepiness; **HQ-CT**, Hyperphagia Questionnaire for use in PWS clinical trials; **MOAS**, Modified Overt Aggression Scale; **SIT**, Self-Injury Trauma scale.
All numeric variables were summarized with median and IQR and compared across groups with a *Wilcoxon Rank Sum Test*.

### Response to GXR

In repeated measures linear regression models, using GEE and an unstructured correlation matrix, there was a decrease in the Aberrant Behavior Checklist score of 9.962 points (95% CI: 19 - 0.8), p=0.03, in subjects receiving GXR treatment as seen in Table 3. A decrease in ABC subscale IV (Hyperactivity) of 5.27 points (95% CI: 1.23 – 9.30), p=0.01, was also observed in this group. While on GXR, subjects demonstrated a reduction in aggressive behaviors as reflected in the decrease in MOAS score of 3.65 points (95% CI: 0.50 – 6.79), p=0.02. The number of skin-picking lesions also decreased in response to GXR, as indicated by the SIT scale scores in the treatment arm; however, this decrease was not significant in comparison to that observed in the placebo arm. There was a highly significant improvement in the overall behavioral symptoms of participants on GXR, as reflected by the lower scores in the psychiatrist-rated CGI-I scales [1.25 points, (95% CI: 0.47 – 2.04), p=0.002]. There were no other statistical differences observed in outcome measures. Fig 2 demonstrates a schematic forest plot of the different effect sizes across measures.

**Fig 2.**
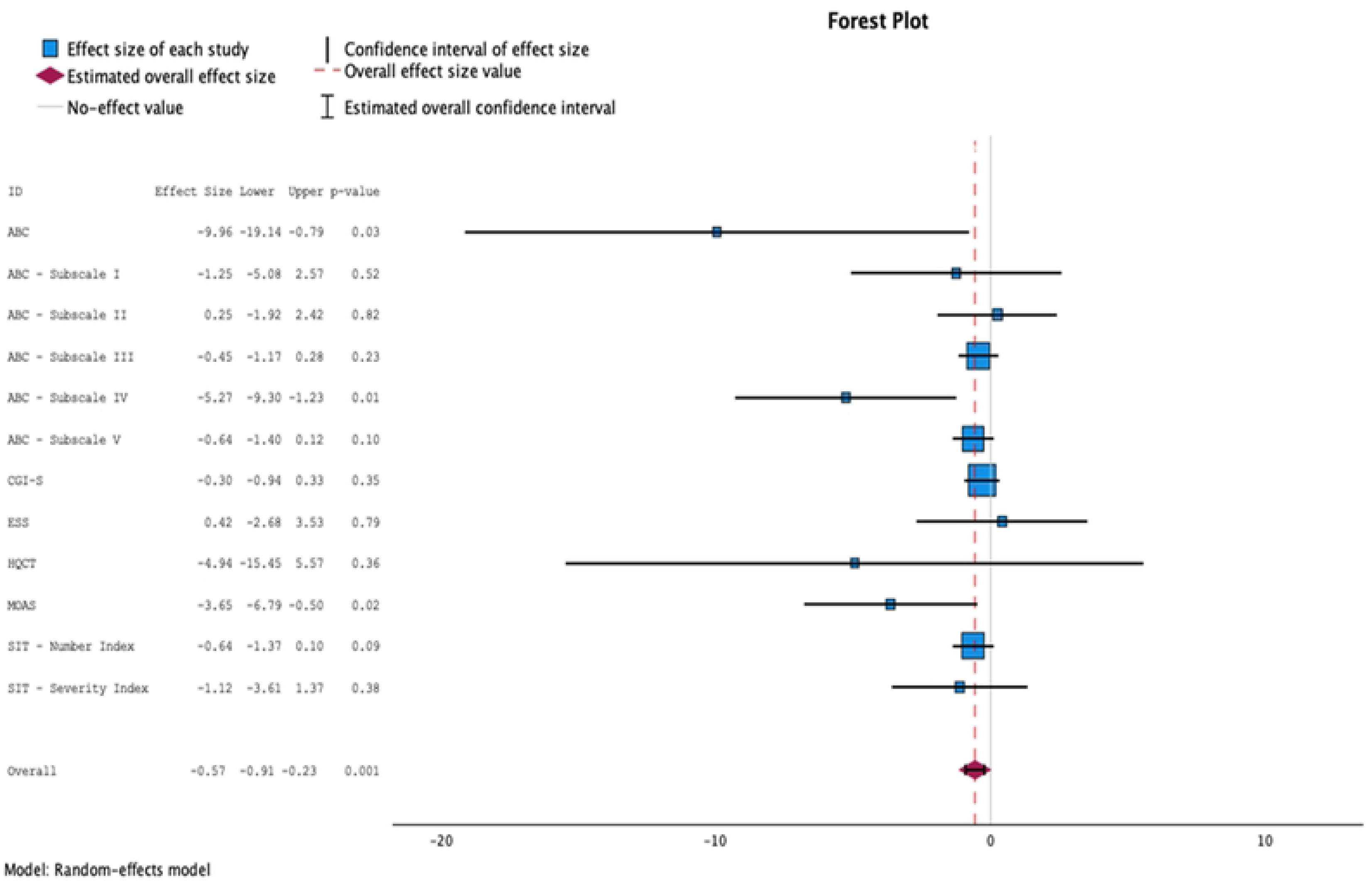
Forest plot. A schematic of the different effect sizes across measures in the two study groups based on GEE Models (Generalized Estimating Equations)

**Table 3.** GEE Models for Additional Outcomes and Scales.

| Outcome Measures | $\beta$ | 95% CI | | P-value |
| --- | --- | --- | --- | --- |
|  |  | LCL | UCL |  |
| ABC | -9.962 | -19.136 | -0.788 | **0.033** |
| ABC - Subscale I, *Irritability* | -1.253 | -5.08 | 2.574 | 0.521 |
| ABC - Subscale II, *Social Withdrawal* | 0.252 | -1.918 | 2.421 | 0.821 |
| ABC - Subscale III, *Stereotypic Behavior* | -0.447 | -1.171 | 0.277 | 0.226 |
| ABC - Subscale IV, *Hyperactivity/Noncompliance* | -5.266 | -9.298 | -1.233 | **0.01** |
| ABC - Subscale V, *Inappropriate Speech* | -0.637 | -1.398 | 0.123 | 0.10 |
| CGI-S | -0.304 | -0.938 | 0.329 | 0.347 |
| CGI-I | -1.25 | -2.036 | -0.465 | **0.002** |
| ESS | 0.425 | -2.684 | 3.533 | 0.789 |
| HQ-CT | -4.94 | -15.45 | 5.57 | 0.357 |
| MOAS (total weighted score) | -3.645 | -6.794 | -0.496 | **0.023** |
| SIT - *Number Index Score* | -0.639 | -1.374 | 0.096 | 0.089 |
| SIT - *Severity Index Score* | -1.121 | -3.611 | 1.368 | 0.377 |

**ABC**, Aberrant Behavior Checklist; **CGI-S**, Clinical Global Impression – Severity scale; **CGI-I**, Clinical Global Impression - Improvement scale; **ESS**, Epworth Sleepiness; **HQ-CT**, Hyperphagia Questionnaire for use in PWS clinical trials; **MOAS**, Modified Overt Aggression Scale; **SIT**, Self-Injury Trauma scale.

### Side Effects and Tolerability

There were no serious adverse events (SAEs) reported by subjects in either of the study groups. However, subjects on GXR were more likely to experience fatigue 29% vs 93%, p = 0.012. Table 4 shows all the treatment-emergent adverse events observed among trial subjects in GXR and placebo groups. There were no other differences in the rate of adverse events amongst the groups.

**Table 4.** Incidence of Adverse Events.

| Adverse Event | Placebo<br>(n=7) | GXR<br>(n=14) | P-value |
| --- | --- | --- | --- |
| Anxiety | 1 (14%) | 2 (14%) | 1.000 |
| Depressed | 1 (14%) | 5 (36%) | 0.615 |
| Dry Mouth | 1 (14%) | 1 (7%) | 0.490 |
| Increased Appetite | 2 (29%) | 3 (21%) | 0.583 |
| Irritable Aggression | 1 (14%) | 5 (36%) | 0.615 |
| Constipation | 0 (0%) | 1 (7%) | 1.000 |
| Fatigue | 2 (29%) | 13 (93%) | **0.012** |
| Enuresis | 1 (14%) | 3 (21%) | 1.000 |
| Excessive Talking | 0 (0%) | 2 (14%) | 1.000 |
| Skin Picking | 0 (0%) | 2 (14%) | 1.000 |
| Diarrhea | 2 (29%) | 1 (7%) | 0.172 |
| Mild Sleep Awakening | 0 (0%) | 3 (21%) | 0.522 |
| Dizziness | 0 (0%) | 1 (7%) | 1.000 |
| Decreased Appetite | 0 (0%) | 1 (7%) | 1.000 |
| Difficult Swallowing | 0 (0%) | 1 (7%) | 1.000 |
| Headache | 1 (14%) | 1 (7%) | 0.490 |
| Increased Energy | 0 (0%) | 1 (7%) | 1.000 |

Adverse events are expressed as frequencies (and percentage) of participants for each treatment-emergent adverse event and compared across groups with a *Fisher Exact Test*.

There were no GXR-related clinically meaningful changes in ECG and QTc-related parameters. No significant changes in vital signs, or lab values were observed between the groups.

The average stable dose of GXR at time of completion of the study was 2.75 mg daily.

## Discussion

Behavioral and psychiatric symptoms are common in the PWS patient population. Individuals with PWS may exhibit impulsive behavior, which can impede daily functioning and lead to risky behavior. ^6, 7, 32^ Furthermore, patients can exhibit aggression and temper tantrums, which leads to significant caregiver burden. ^5, 33^ Skin-picking is another behavioral symptom commonly seen in this patient population, which may get complicated with infections and scarring at the site of damage. ^19^ The overall burden of illness over the lifetime of an individual with PWS is compounded by these behavioral problems.

Currently, the commonly used pharmacological treatment options for behavioral management in individuals with PWS are SRIs, stimulants, and atypical antipsychotics. Their use in PWS is widespread despite the lack of empirical placebo-controlled studies to demonstrate efficacy. These medications are associated with significant risks including weight gain for antipsychotics, psychosis for SRIs, and increased skin excoriation with stimulants. With the exception of NAC, options are limited for the treatment of skin-picking behavior. ^19, 20, 21, 1, 34^ Therefore, investigating relatively safe medication options that can effectively manage several impairing behavioral symptoms seen in PWS is warranted to positively impact the lives of patients and to reduce caregiver fatigue.

This randomized double-blind, placebo-controlled, fixed-flexible dose clinical trial with open-label continuation, was designed to explore the efficacy, safety, and tolerability of guanfacine extended release in the management of aggression and self-injurious behavior (skin picking) in individuals with PWS. The results of this study demonstrate that GXR ameliorates overall behavioral disturbances as measured by the ABC scale. GXR was particularly effective against hyperactivity in PWS. This finding is expected since GXR has established evidence for the treatment of ADHD in the general population. ^35, 36^ Additionally, GXR led to a significant reduction in aggressive behavior as measured by the MOAS. Our study also showed a positive response to GXR in terms of skin picking, with a reduction in the number of lesions in the treatment group, although it was not statistically significant as compared to that in the placebo group. Notably, subjects on GXR showed a significant improvement in their overall presentation as measured by the CGI-I scale.

GXR was generally well tolerated with the most commonly reported side effect being fatigue, commensurate with the findings in the general population. ^37^ It is important to note that despite this, the overall level of daytime sleepiness as measured by the ESS did not show any statistical difference from placebo. This finding is of particular salience given the high rates of daytime sleepiness experienced in this population. ^38^ There were no serious adverse events reported in the GXR group. Another key factor to consider in this study is that, none of the subjects exposed to GXR experienced weight gain. There was no difference noted between the GXR and placebo groups with respect to ECG changes, blood pressure, or heart rate.

Several outcomes trended towards a positive response but failed to reach a statistical significance level. In particular, a positive signal was observed for hyperphagia and skin picking. These need to be confirmed in future studies with a larger sample size.

Individuals with PWS suffer from growth hormone (GH) deficiency, which contributes to the short stature and the central obesity. GH replacement therapy has shown beneficial effects in aiding linear growth, and restoring normal body composition, bone density, as well as lipid profile values in individuals with PWS. ^39 40^ At the time of this writing, GH is the only medication that has an FDA approved indication for the management of PWS. ^41^ Clonidine is an α2-adrenoceptor agonist similar to GXR, and both medications have been shown to increase GH levels in a dose-dependent manner. However, GXR is less sedating compared to clonidine and may be a better alternative in this setting. ^42^ Although not measured in the current study, the possible physical benefits of increased GH could indicate an additional advantage of GXR in comparison to other medications used for the behavioral management of individuals with PWS. This could be further elucidated in prospective trials.

### Limitations

The multiple limitations of this study need to be highlighted. Although there were no gender differences between the active and placebo arms, the majority of patients enrolled were male. A larger study could reduce the possibility of there being a gender based moderating effect on the results. A larger sample size would also allow stratification of the results by PWS genotype. Similarly, the participants were predominantly Caucasian hence limiting generalizability of findings. The study was conducted at a single site, limiting the diversity in participants that can be achieved from a multicenter study. It is challenging to establish external validity in a single-site study, due to limitations of accounting for the variance in environmental and practitioner-based factors. Another factor that must be considered is the relatively high prevalence of ADHD and autism in PWS.^6^ Therefore, there is a possibility that the observed effects may be moderated by these co-existing conditions.

Despite these challenges to the study, the results of this clinical trial, along with prior knowledge of the side effects posed by the alternatives available to practitioners, suggests that GXR should be the first line treatment for behavioral problems, specifically aggression, irritability, and hyperactivity in individuals with PWS.

## Conclusions

This randomized, fixed-flexible dose placebo-controlled clinical trial, with open label extension, aimed to provide a supporting framework for the utilization of GXR in the management of behavioral symptoms comorbid with PWS. The assessed symptoms included aggression, irritability, inattention/hyperactivity, and skin-picking. GXR treatment leads to global improvement in symptoms, particularly those related to behavioral disturbance in persons with PWS. Specifically, GXR reduces aggression and hyperactivity in this population. Additionally, there are possible beneficial effects on skin picking behavior. The most commonly reported side effect was fatigue, but overall daytime sleepiness was not worse than placebo. There were no serious adverse effects reported. GXR did not lead to any change in weight, heart rate, blood pressure, or cardiac rhythm, as compared to placebo. The results of this clinical trial suggests that GXR is an effective and relatively safe treatment modality for behavioral problems in patients with PWS.

### Clinical Significance

Individuals with PWS often present with challenging behavioral disturbances such as aggression/agitation, skin-picking, and ADHD symptoms like impulsivity and inattention. Current management strategies for behavioral disturbances in PWS patients show limited efficacy and can have significant side effects. Although other studies have been conducted to assess treatment options against hyperphagia, this is the first randomized placebo-controlled trial examining the efficacy of a psychotropic medication against the behavioral manifestations of PWS. This randomized placebo-controlled clinical trial demonstrated that GXR is effective in the management of behavioral disturbances in PWS while being well tolerated. The authors recommend that GXR be considered as a first line treatment strategy for the management of behavioral problems, especially aggression and hyperactivity, associated with PWS.

## Data Availability

All relevant data are within the manuscript and its Supporting Information files and will be available upon its publication.

## Supporting Information

**S1 Checklist. CONSORT Checklist**

## Acknowledgments

The authors would like to thank our hospital pharmacists Jason Brady and Rina Evans for their invaluable contribution. We would also like to thank our clinical research coordinators Otuwe Anya and Hasan Mustafic, without whom this study would not have been possible.

